# Effectiveness and Safety of Propofol at Low Doses for Emergency Department Treatment of Migraine

**DOI:** 10.1101/2024.10.09.24315176

**Authors:** Stéphane Côté, David Simonyan, Myriam Mallet, Simon Baril, Laurie Ouellet, Simon Berthelot

## Abstract

**Introduction:** A migraine treatment protocol implemented in the emergency department of an urban hospital allowed us to evaluate the effectiveness of propofol compared to metoclopramide as well as the safety of the protocol.

**Methods:** We reviewed the health records of all patients aged 16 years and older treated with propofol for migraine between May 2014 and August 2017 at a teaching hospital in Québec City (CHUL). The care protocol consisted of administering propofol (20 mg) every 5–10 minutes as needed (up to 6 doses), monitoring vital signs before and after each dose and continuous cardiac monitoring. The primary outcome measure was the mean reduction of pain following first-line therapy (propofol or metoclopramide). The secondary outcome measures were 1) adjusted relative risks of requiring rescue medication after first-line therapy; 2) incidence of the following side effects of propofol received as first or second-line therapy: low arterial pressure (< 90 systolic or < 65 mean), desaturation, excessive sedation, arrhythmia. The cohorts were paired for gender, age, triage priority, and month/year of ED visit.

**Results:** Files of 34 patients given propofol and 58 given metoclopramide as first-line treatment were analyzed. Five metoclopramide-treated patients received propofol as rescue medication. Among propofol-treated patients, 29.4% experienced pain relief compared to 66% in the metoclopramide group (p < 0.001). Rescue medication was more frequent in first-line propofol patients (82.4% versus 37.9%, p < 0.001). In this group, four participants (10.3%) received intravenous fluid bolus for mean blood pressure below 60, but no persistent desaturation, bradycardia, excessive sedation, or arrhythmia was recorded.

**Conclusion:** Though less effective than metoclopramide, propofol at low doses may be an alternative to treat migraine in the ED. Monitoring of vital signs (especially blood pressure) would be prudent but continuous nursing is likely unnecessary.

## INTRODUCTION

Migraine is a common cause of emergency department (ED) visits (1). Current best estimates indicate that the global prevalence of migraine is 14-15% and represents 4.9% of years lived with disability worldwide (2). Migraine patients presenting to the ED have often experienced therapeutic failure (5). A safe and effective pharmacological alternative for quick relief from migraine pain is often difficult to identify in the ED.

The targets of medication are often multiple and complex, and numerous options have been used to treat acute migraine. The most common of these include nonsteroidal anti-inflammatory agents, neuroleptics, dopamine-antagonist (e.g., metoclopramide) and triptans (6). Reports of the effectiveness of subanesthetic doses of propofol began to appear more than 20 years ago (7-14). Although the exact pharmacological mechanism of propofol as an anti-migraine is unclear, agonism at gamma-aminobutyric acid A (GABAa) receptors is likely involved, producing anxiolytic, sedative, and anesthetic effects. GABA receptor agonism affects calcium channels and may also inhibit N-methyl-D-aspartate (NMDA) receptors, leading to neuronal hyperpolarization and inhibition of the neuronal firing that might lead to migraine (7, 15). One of the challenges of using propofol is its significant side-effect profile, which includes arterial hypotension and respiratory depression (16, 17). Although low doses are used to treat migraine, the patients need more monitoring than with other alternatives, since the safety of propofol remains to be demonstrated unequivocally for this indication.

The aim of this study was 1) to compare the effectiveness of propofol with that of metoclopramide and 2) to assess its safety for the treatment of migraines in the ED.

## METHODS

### Study design and setting

We conducted a health records review of patients who received intravenous propofol as a first-line or second-line ED treatment for migraine at the CHUL (a teaching hospital in Québec City, Canada, receiving 78,000 visits annually) between May 2014 and August 2017. Patients were identified through a mandatory specific prescription form that emergency physicians had to fill when using propofol for this off-label indication. For comparison purposes, we also extracted clinical data from a cohort of migraine patients who received metoclopramide initially. The two cohorts were paired for sex, age, triage priority, and month/year of the ED visit. This study received approval from the CHU de Québec-Université Laval research ethics board (authorization no. 2017-3012).

### Participants

We reviewed and included all charts of patients who received propofol during the study period if they were at least 16 years old and treated for migraine without aura or with typical aura as defined in the International Headache Society (IHS) classification (18). Excluded from the analysis were patients treated with propofol for headaches not meeting the IHS criteria and those who received propofol despite the presence of contraindications identified in the local care protocol, namely pregnancy, severe coronary heart disease (ASA ≥3/4), heart failure, severe chronic pulmonary disease (FEV_1_ <50%), familial dyslipidemia, neuromuscular disease, oxygen dependence, hemodynamic instability, allergies to egg or soy.

### Therapeutic guide

A therapeutic guide implemented in May 2014 for using propofol to treat migraine specifies administering 20 mg (intravenously) every 5 to 10 minutes as needed, up to 6 doses (see web-appendix). Vital signs and pain assessment are recorded before and after each dose. Cardiac and saturation monitoring is continuous. A bedside nurse assesses the patient’s pain score, state of consciousness and hemodynamic parameters throughout administration of the drug. Patients are considered relieved, and administration is stopped, when the pain score decreases to 2 or less on a scale from 0 (no pain) to 10 (worst pain).

### Data sources and collection

All patient electronic records were extracted by two independent evaluators, and disagreements with the charts were resolved by discussion. A third evaluator participated when the discussion did not reach a consensus. Demographic and clinical characteristics were collected on a standardized Excel spreadsheet and included the following variables: age, biological sex, time, date and month of ED visit, comorbidities, regular medication, vital signs, pain assessment (10-level) and sedation on the 4-level Pasero scale (19) from 1 (easy to rouse) to 4 (unresponsive) on ED arrival and discharge, pre- and post-administration of propofol, use of co-analgesics or rescue medication. Since metoclopramide administration was not standardized in a protocol, pain assessment was not translated systematically to the 10-level scale when the patient was discharged. When no final pain score was recorded on the chart of a patient discharged after treatment with metoclopramide, we extrapolated a score from the nurse’s discharge note, assigning a score of 2 if partial or almost complete pain relief was mentioned, and 0 if the nurse specifically indicated that the patient had no pain at discharge.

### Outcomes

The primary outcome measure was the mean reduction of pain after first-line treatment (propofol or metoclopramide). The secondary outcomes measures were 1) adjusted relative risk of requiring rescue medication; 2) incidence of the following side effects for patients who received propofol (as first-line therapy or rescue medication): low arterial pressure (< 90 systolic or < 65 mean), desaturation (SaO_2_ < 92%), excessive sedation (Pasero scores of 3 or 4), and any arrhythmia.

### Statistical analysis

Patient characteristics are presented as means, counts, and proportions. Mean pain scores pre- and post-propofol (first- and second-line treatments) were compared using a generalized linear regression model (estimating equations) designed for repeated measures. Incidences of adverse effects are reported as proportions with 95% confidence intervals. We also compared propofol to metoclopramide using Student’s t-test on the mean differences of pre- and post-treatment pain scores. A Chi-squared test was conducted to compare the proportions of each group that needed rescue medication. The adjusted relative risk was estimated for this need using a log-binomial regression model and a propensity score generated using a logistic regression model with treatment groups (propofol versus metoclopramide) as the dependent variable. The independent variables were age, sex, anti-migraine medication taken before arriving at the ED, triage score, chronic obstructive pulmonary disease, chronic use of antihypertensives or anxiolytics, and clinical parameters upon arrival in the ED including pain level, oxygen saturation, systolic and diastolic blood pressure. All analyses were performed with SAS version 9.4 (SAS Institute, Cary, NC, USA) with a two-sided significance level set at p < 0.05.

## RESULTS

The demographics and other basic characteristics of the participants are summarized in Table 1. Over the 3-year study period, 34 participants received propofol as first-line treatment in accordance with the therapeutic guide. They were compared with 58 participants who received metoclopramide as first-line treatment. Among these, 5 received propofol as rescue medication. They are described separately in Table 1. There were no significant differences in age, sex, or co-morbidity between the propofol and metoclopramide first-line groups. The baseline vital signs and pain scores at ED triage were also comparable. However, 12.7% more participants in the metoclopramide group had taken anti-migraine medication before arriving at the ED.

**TABLE 1.** Characteristics of the participants ^a^.

| Characteristic | Propofol<br>1<sup>st</sup> line (n = 34) | Propofol<br>2<sup>nd</sup> line<sup>b</sup> (n = 5) | Metoclopramide<br>(n = 53) |
| --- | --- | --- | --- |
| Mean age in years (SD)<sup>c</sup> | 37.1 (14.7) | 45.6 (14.7) | 34.8 (12.4) |
| Female, n (%) | 27 (79.4) | 4 (80.0) | 43 (81.1) |
| Canadian Triage and Acuity<br>Score |  |  |  |
| 2 | 6 (17.6) | 1 (20.0) | 9 (17.0) |
| 3 | 24 (70.6) | 4 (80.0) | 41 (77.3) |
| 4 | 4 (11.8) | 0 (0) | 3 (5.7) |
| Mean vital signs at triage<br>(SD) |  |  |  |
| Systolic blood pressure | 130.3 (17.3) | 126.6 (17.6) | 132.1 (19.0) |
| Diastolic blood pressure | 82.2 (11.8) | 83.8 (8.0) | 83.7 (10.8) |
| Oxygen saturation | 99.1 (1.3) | 99.3 (1.5) | 98.7 (1.5) |
| Respiratory rate | 18.6 (4.7) | 17 (1.4) | 17.7 (2.6) |
| Comorbidities |  |  |  |
| Hypertension | 2 (5.9) | 0 (0) | 6 (11.3) |
| Coronary artery disease | 0 (0) | 0 (0) | 2 (3.8) |
| Asthma/Chronic<br>pulmonary disease | 3 (8.8) | 1 (20.0) | 10 (18.9) |
| Regular medication |  |  |  |
| Opioids | 0 (0) | 0 (0) | 2 (3.8) |
| Benzodiazepines | 0 (0) | 1 (20.0) | 3 (5.7) |
| Antihypertensives | 4 (11.8) | 0 (0) | 5 (9.4) |
| Antimigraine taken prior to<br>arrival at the ED | 22 (64.7) | 3 (60.0) | 41 (77.4) |
| Received concomitant<br>medication, n (%)<sup>d</sup> | 4 (11.8) | 4 (80.0) | 43 (81.1) |
| Acetaminophen | 1 (2.9) | 1 (20.0) | 14 (26.4) |
| Naproxen | 2 (5.9) | 2 (40.0) | 16 (30.2) |
| Dexamethasone | 2 (5.9) | 1 (20.0) | 5 (9.4) |
| Lorazepam | 0 (0) | 1 (20.0) | 4 (7.5) |
| Diphenhydramine | 0 (0) | 1 (20.0) | 21 (39.6) |
| Opioids | 0 (0) | 0 (0) | 0 (0) |

<sup>a</sup> All data are reported as n (%), unless otherwise indicated.
<sup>b</sup> All patients who received propofol as 2nd line medication received metoclopramide as their first antimigraine agent in the ED and are therefore included in the metoclopramide group.
<sup>c</sup> Standard deviation
<sup>d</sup> Given at the same time as propofol and metoclopramide.

As shown in Figure 1 and Table 2, repeated doses of propofol (first or second line) decreased pain intensity significantly up to the sixth dose. However, only 29.4% of patients given first-line treatment with propofol achieved a pain level of 2 or less, compared to 66% of patients at comparable pain levels upon arrival in the ED given metoclopramide (p < 0.001). However, 81.1% of these patients received co-analgesic medications, compared to only 11.8% of patients treated with propofol.

**Figure 1.**
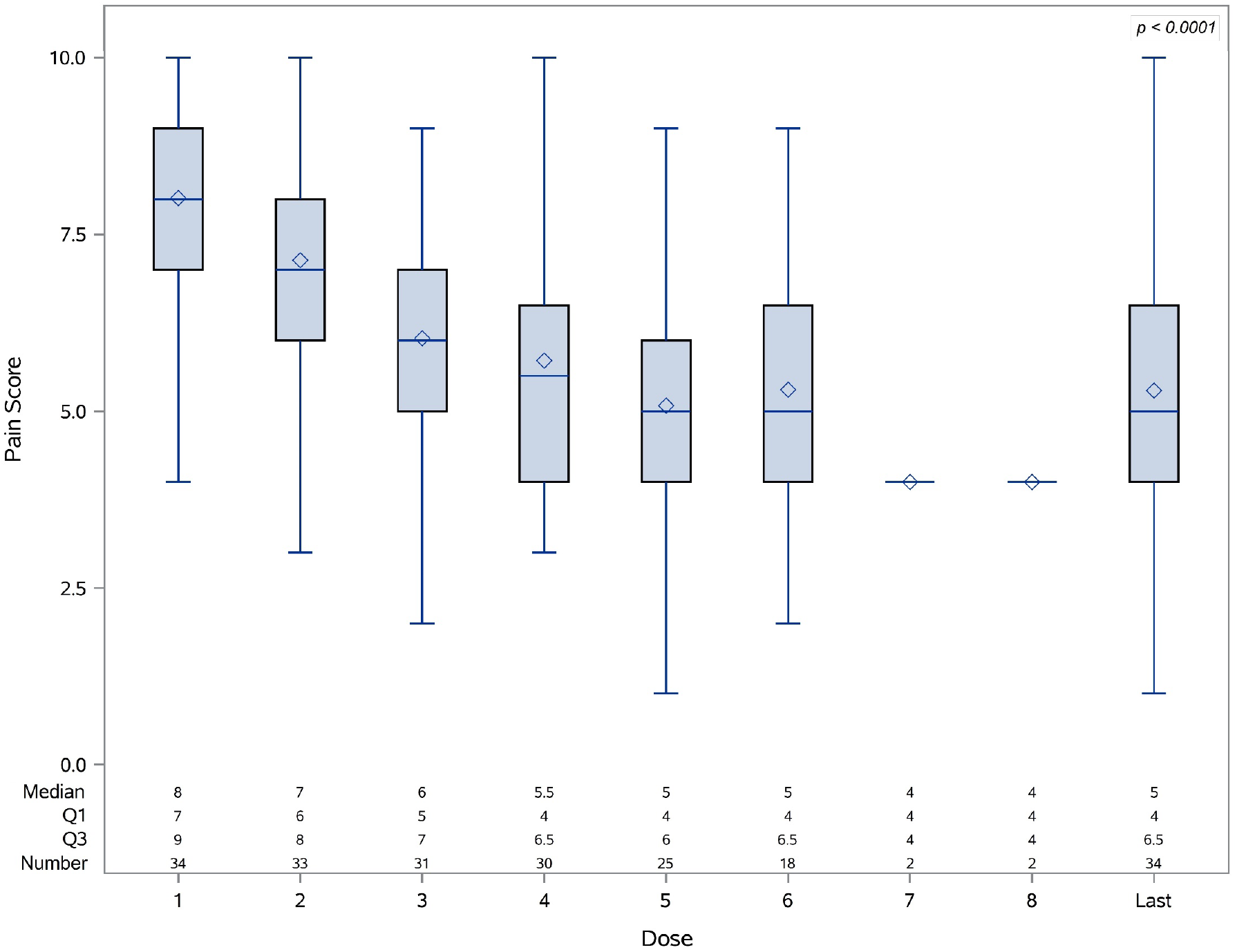
Box plot of migraine pain intensity recorded following repeated low doses of propofol as a first-line therapy (n = 34)

**Table 2.** Mean pain score recorded before and after first-line treatment with propofol or metoclopramide.

| Measurement | Propofol<br>(n = 34) | Metoclopramide<br>(n = 58) | *P value*<sup>a</sup> |
| --- | --- | --- | --- |
| Initial pain score<sup>b</sup> | 8.2 (7.6-8.7)<sup>c</sup> | 7.7 (7.2-8.2) | 0.21 |
| Final pain score | 4.3 (3.3-5.2) | 2.1 (1.3-2.9) | < 0.001 |
| Before-after difference | 3.8 (2.9-4.8) | 5.6 (4.8-6.4) | 0.006 |
| Length of ED stay<br>(hours) | 7.0 (5.2-8.9) | 7.5 (5.2-9.8) | 0.78 |
| Percent of patients<br>relieved of pain (score ≤<br>2) | 29.4 | 66 | < 0.001 |

<sup>a</sup> Based on Student's t-test
<sup>b</sup> From 0 (no pain) to 10 (worst pain)
<sup>c</sup> All results are reported as means with 95% confidence intervals, unless otherwise indicated

The proportion of patients who required a rescue medication was higher in the propofol group (82.4% vs 37.9%, p < 0.001, Table 3). The adjusted risk of requiring rescue medication was 81% higher for this group. In 67% of rescues in the propofol group, the medication used was metoclopramide, whereas propofol was used in only 8.6% of the metoclopramide group needing rescue.

**TABLE 3.**
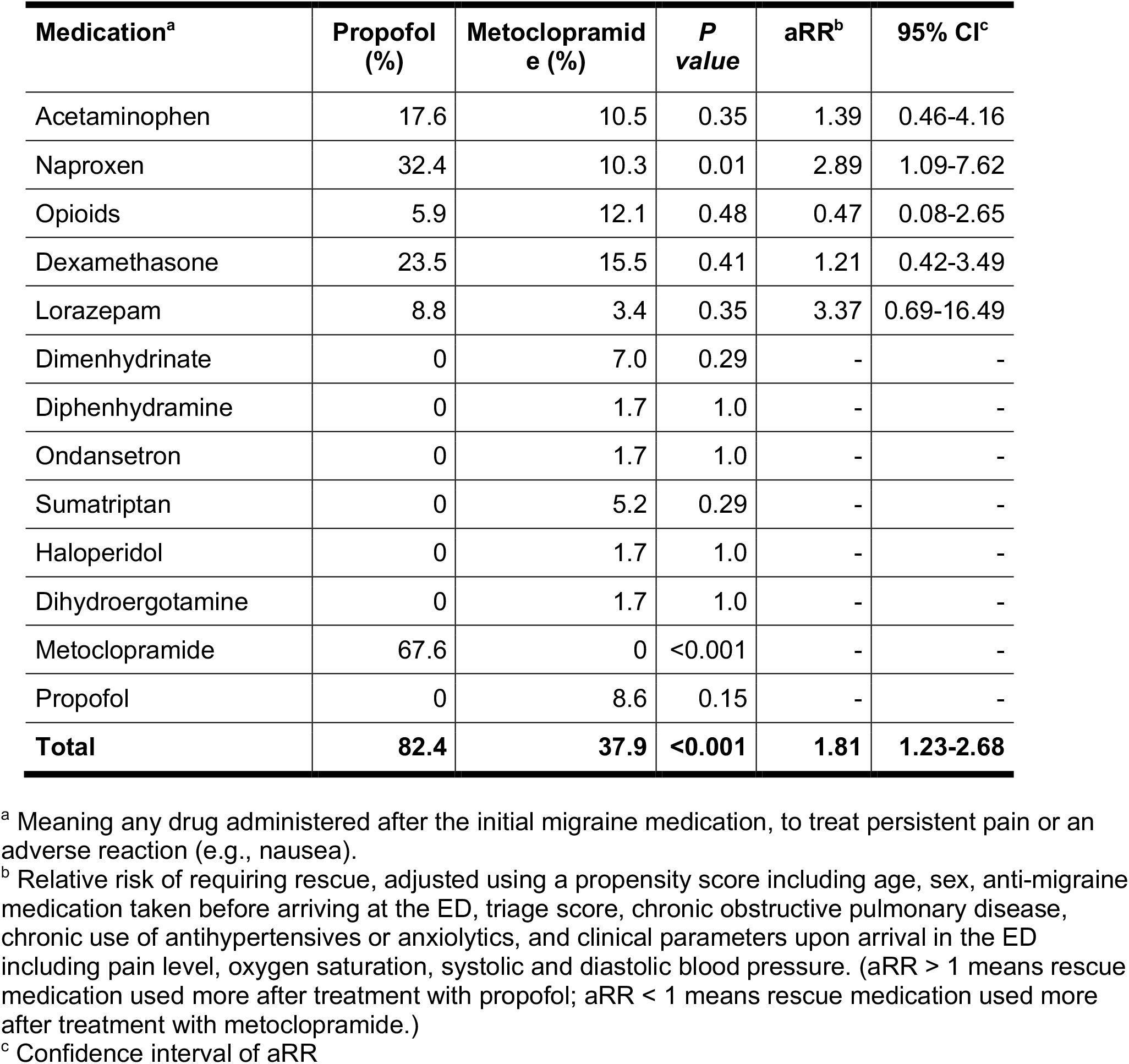
Rescue medications used after ED treatment of migraine with propofol (n = 34) or metoclopramide (n = 58)

| **Medication<sup>a</sup>** | **Propofol (%)** | **Metoclopramide (%)** | ***P* value** | **aRR<sup>b</sup>** | **95% CI<sup>c</sup>** |
| --- | --- | --- | --- | --- | --- |
| Acetaminophen | 17.6 | 10.5 | 0.35 | 1.39 | 0.46-4.16 |
| Naproxen | 32.4 | 10.3 | 0.01 | 2.89 | 1.09-7.62 |
| Opioids | 5.9 | 12.1 | 0.48 | 0.47 | 0.08-2.65 |
| Dexamethasone | 23.5 | 15.5 | 0.41 | 1.21 | 0.42-3.49 |
| Lorazepam | 8.8 | 3.4 | 0.35 | 3.37 | 0.69-16.49 |
| Dimenhydrinate | 0 | 7.0 | 0.29 | - | - |
| Diphenhydramine | 0 | 1.7 | 1.0 | - | - |
| Ondansetron | 0 | 1.7 | 1.0 | - | - |
| Sumatriptan | 0 | 5.2 | 0.29 | - | - |
| Haloperidol | 0 | 1.7 | 1.0 | - | - |
| Dihydroergotamine | 0 | 1.7 | 1.0 | - | - |
| Metoclopramide | 67.6 | 0 | <0.001 | - | - |
| Propofol | 0 | 8.6 | 0.15 | - | - |
| **Total** | **82.4** | **37.9** | **<0.001** | **1.81** | **1.23-2.68** |

<sup>a</sup> Meaning any drug administered after the initial migraine medication, to treat persistent pain or an adverse reaction (e.g., nausea).
<sup>b</sup> Relative risk of requiring rescue, adjusted using a propensity score including age, sex, anti-migraine medication taken before arriving at the ED, triage score, chronic obstructive pulmonary disease, chronic use of antihypertensives or anxiolytics, and clinical parameters upon arrival in the ED including pain level, oxygen saturation, systolic and diastolic blood pressure. (aRR > 1 means rescue medication used more after treatment with propofol; aRR < 1 means rescue medication used more after treatment with metoclopramide.)
<sup>c</sup> Confidence interval of aRR

Among recipients of propofol as a first line treatment (n = 34) or as a rescue medication (n = 5), no significant or persistent desaturation (SaO_2_ < 92%), bradycardia (heart rate < 60) or excessive sedation (Pasero score 3 or 4) were recorded. At the end of first-line treatment, 54.5% of the participants had a transient decrease in their mean blood pressure to below 60 mmHg, and 6.1% had a systolic blood pressure below 90 mmHg. Intravenous fluid bolus was undertaken for four participants in the propofol group (10.3%), but no vasopressor was needed.

## DISCUSSION

This review of health records shows that repeated low doses of propofol are effective at reducing pain in patients presenting with migraines. No desaturation or decreased level of consciousness occurred, although 10% of patients received a fluid bolus for minor drops in blood pressure. However, metoclopramide appears to be more effective in the ED setting: more patients felt their pain reduced to 2 or lower, and fewer required rescue medication. Nevertheless, propofol administered at low doses appears to be a potential alternative for this indication, requiring only blood pressure monitoring to ensure patient safety.

### Previous literature

The ED use of propofol to treat migraines has been reviewed systematically twice in recent years. The first of these (20) considered 9 studies that covered 290 patients who received propofol as first-line treatment: 5 case reports or case series, 1 retrospective cohort study, and 3 randomized controlled studies. The subsequent review (21) focused on the effectiveness of parenteral agents. We also examined 4 randomized trials (22-25) of propofol, including one with metoclopramide or granisetron as a co-intervention for nausea (24). Despite the design problems of reviews based on health records, our study included a cohort of 92 patients, comparable in size to the largest randomized studies to date (22, 26). Although propofol was found not superior to placebo in a cohort study of 40 patients (25), all other reports indicate that this molecule is effective for the treatment of migraine. We found that 29.4% of patients given propofol as first-line treatment were relieved completely of pain (score ≤ 2) and a mean pain reduction of 46.3% was obtained. This diverges from studies in which 84.4% of the participants were relieved of pain (22) or the mean migraine pain score was reduced by 87.5% (26). The lower effectiveness observed in our study may be due to differences in the cohorts and protocols. Only 22% of our propofol group had received medication before admission to the ED, versus 93.3% (22). Furthermore, we administered propofol in iv doses of 20 mg for a total of 120 mg, versus an initial 30-40 mg subcutaneous dose followed by boluses of 10-20 mg up to a total of 120 mg. Finally, we considered that the patient was relieved of pain when the score reached 2 or less on the 10-level scale, versus defining responsiveness to therapy as at least a 4-point reduction in VAS.

The other point to consider when using propofol is its safety. The adverse effects of propofol (cardiovascular, e.g., bradycardia, hypotension; respiratory, e.g., ventilatory depressant, desaturation, hypoxemia, apnea) are well documented and controlled with constant monitoring by qualified medical personnel (16, 17). Since typical ED staff regard with apprehension the use of propofol outside intensive care units or resuscitation rooms, we implemented a strict protocol to evaluate side effects. Our results confirm previous reports that using propofol at subanesthetic doses causes very few side effects (20, 21). The safety profile of low dose propofol in the ED for treatment of migraine thus appears favorable.

### Clinical implications

This study confirms previous findings that propofol may be considered a safe alternative for treatment of migraine in the ED. However, given the well-demonstrated effectiveness of metoclopramide, this agent may be the better first-line option. The care protocol prompting this study included very close monitoring of patients with nearly continuous nursing presence. The few side effects recorded demonstrate that propofol is safe, and at doses much lower than those used for procedural sedation or intubation, blood pressure monitoring is likely sufficient precaution. Our data suggest that propofol is safe enough to be used without constant bedside nursing.

### Research implications

The effectiveness of propofol for the treatment of migraine is not negligible but the scientific data available remain insufficient to justify its addition to the list of standard therapeutic options. Randomized controlled studies with larger sample sizes are needed to obtain a reliable estimation of its true effectiveness (27, 28). Furthermore, the doses of propofol currently used are chosen empirically and often differ from one study to another, which may explain its variable efficacy. A protocol must be developed to establish a pharmacokinetic dose-response curve (EC50). Patient weight-matched doses of propofol will help answer the questions that remain.

### Limitations and strengths

A study based on review of health records has inherent limitations that should be considered when interpreting the results. One possible confounding factor in the present study is that the clinicians in the metoclopramide group were not required to follow a standardized protocol. Side effects such as extra-pyramidal symptoms were not recorded, and since the pain score was not always noted at discharge from the ED, we presumed a score of 2 or less, based on physician and nurse notes. The strength of our study lies in the rigorous supervision of a propofol administration protocol that we had established, which ensured data gathering throughout the dosing period. Two reliable indicators emerge from our study. First, we were able to measure the need for rescue medication in both groups, since all administering of medication in the hospital is fully recorded. Second, the patients treated with propofol benefited from close monitoring of vital signs. This makes us confident in our statement that no significant adverse effects occurred.

## CONCLUSION

In this study, we evaluated the effectiveness and safety of administering propofol to treat migraines in the ED. The results suggest that propofol can be a safe alternative as a second-line option after the administration of neuroleptics, primarily metoclopramide, which appears to be the first choice in many EDs. If propofol is administered, monitoring of vital signs, especially blood pressure, appears prudent, but continuous bedside nursing is likely unnecessary.

## Data Availability

All data produced in the present study are available upon reasonable request to the authors

## REFERENCES

1. Friedman BW, Hochberg ML, Esses D, Grosberg B, Corbo J, Toosi B, et al. Applying the International Classification of Headache Disorders to the emergency department: an assessment of reproducibility and the frequency with which a unique diagnosis can be assigned to every acute headache presentation. Ann Emerg Med. 2007;49(4):409–19, 19 e1-9.

2. Steiner TJ, Stovner LJ. Global epidemiology of migraine and its implications for public health and health policy. Nat Rev Neurol. 2023;19(2):109–17.

3. Doretti A, Shestaritc I, Ungaro D, Lee JI, Lymperopoulos L, Kokoti L, et al. Headaches in the emergency department -a survey of patients’ characteristics, facts and needs. J Headache Pain. 2019;20(1):100.

4. Hazard E, Munakata J, Bigal ME, Rupnow MF, Lipton RB. The burden of migraine in the United States: current and emerging perspectives on disease management and economic analysis. Value Health. 2009;12(1):55–64.

5. Gelfand AA, Goadsby PJ. A Neurologist’s Guide to Acute Migraine Therapy in the Emergency Room. Neurohospitalist. 2012;2(2):51–9.

6. Wells S, Stiell IG, Vishnyakova E, Lun R, Nemnom MJ, Perry JJ. Optimal management strategies for primary headache in the emergency department. CJEM. 2021;23(6):802–11.

7. Krusz JC, Scott V, Belanger J. Intravenous propofol: unique effectiveness in treating intractable migraine. Headache. 2000;40(3):224–30.

8. Giampetro D, Ruiz-Velasco V, Pruett A, Wicklund M, Knipe R. The Effect of Propofol on Chronic Headaches in Patients Undergoing Endoscopy. Pain research & management. 2018;2018:6018404.

9. Simmonds MK, Rashiq S, Sobolev IA, Dick BD, Gray DP, Stewart BJ, et al. The effect of single-dose propofol injection on pain and quality of life in chronic daily headache: a randomized, double-blind, controlled trial. Anesthesia and analgesia. 2009;109(6):1972–80.

10. Bloomstone JA. Propofol: a novel treatment for breaking migraine headache. Anesthesiology. 2007;106(2):405–6.

11. Udelsmann A, Saccomani P, Dreyer E, da Costa AL. Treatment of status migrainosus by general anesthesia: a case report. Braz J Anesthesiol. 2015;65(5):407–10.

12. Drummond-Lewis J, Scher C. Propofol: a new treatment strategy for refractory migraine headache. Pain Med. 2002;3(4):366–9.

13. Mendes PM, Silberstein SD, Young WB, Rozen TD, Paolone MF. Intravenous propofol in the treatment of refractory headache. Headache. 2002;42(7):638–41.

14. Thurlow JA. Hemiplegia following general anaesthesia: an unusual presentation of migraine. Eur J Anaesthesiol. 1998;15(5):610–2.

15. Dhir A, Lossin C, Rogawski MA. Propofol hemisuccinate suppresses cortical spreading depression. Neurosci Lett. 2012;514(1):67–70.

16. Sahinovic MM, Struys M, Absalom AR. Clinical Pharmacokinetics and Pharmacodynamics of Propofol. Clin Pharmacokinet. 2018;57(12):1539–58.

17. Marik PE. Propofol: therapeutic indications and side-effects. Curr Pharm Des. 2004;10(29):3639–49.

18. Headache Classification Committee of the International Headache Society (IHS) The International Classification of Headache Disorders, 3rd edition. Cephalalgia. 2018;38(1):1–211.

19. Pasero C. Assessment of sedation during opioid administration for pain management. J Perianesth Nurs. 2009;24(3):186–90.

20. Piatka C, Beckett RD. Propofol for Treatment of Acute Migraine in the Emergency Department: A Systematic Review. Acad Emerg Med. 2020;27(2):148–60.

21. Kirkland SW, Visser L, Meyer J, Junqueira DR, Campbell S, Villa-Roel C, et al. The effectiveness of parenteral agents for pain reduction in patients with migraine presenting to emergency settings: A systematic review and network analysis. Headache. 2024;64(4):424–47.

22. Moshtaghion H, Heiranizadeh N, Rahimdel A, Esmaeili A, Hashemian H, Hekmatimoghaddam S. The Efficacy of Propofol vs. Subcutaneous Sumatriptan for Treatment of Acute Migraine Headaches in the Emergency Department: A Double-Blinded Clinical Trial. Pain practice: the official journal of World Institute of Pain. 2015;15(8):701–5.

23. Mitra B, Roman C, Mercier E, Moloney J, Yip G, Khullar K, et al. Propofol for migraine in the emergency department: A pilot randomised controlled trial. Emergency medicine Australasia: EMA. 2020;32(4):542–7.

24. Abiri S, Chegin M, Soleimani R, Hatami N, Kalani N, Rayatdoost E. Propofol + Granisetron vs. Propofol + Metoclopramide in Symptom Management of Acute Migraine Headache; a Double-Blind Randomized Clinical Trial. Arch Acad Emerg Med. 2022;10(1):e19.

25. Meek R, Graudins A, McDonald M, McGannon D, Limm E. Comparing propofol with placebo for early resolution of acute migraine in adult emergency department patients: A double-blind randomised controlled trial. Emergency medicine Australasia: EMA. 2021;33(3):465–72.

26. Soleimanpour H, Ghafouri RR, Taheraghdam A, Aghamohammadi D, Negargar S, Golzari SE, et al. Effectiveness of intravenous dexamethasone versus propofol for pain relief in the migraine headache: a prospective double blind randomized clinical trial. BMC Neurol. 2012;12:114.

27. Orr SL, Aube M, Becker WJ, Davenport WJ, Dilli E, Dodick D, et al. Canadian Headache Society systematic review and recommendations on the treatment of migraine pain in emergency settings. Cephalalgia. 2015;35(3):271–84.

28. Abdelmonem H, Abdelhay HM, Abdelwadoud GT, Alhosini ANM, Ahmed AE, Mohamed SW, et al. The efficacy and safety of metoclopramide in relieving acute migraine attacks compared with other anti-migraine drugs: a systematic review and network meta-analysis of randomized controlled trials. BMC Neurol. 2023;23(1):221.

